# Discontinuation and Reinitiation of GLP-1 Receptor Agonists Among US Adults with Overweight or Obesity

**DOI:** 10.1101/2024.07.26.24311058

**Authors:** Patricia J. Rodriguez, Vincent Zhang, Samuel Gratzl, Duy Do, Brianna Goodwin Cartwright, Charlotte Baker, Ty J. Gluckman, Nicholas Stucky, Ezekiel J. Emanuel

## Abstract

**Importance:** Adherence to GLP-1 RA is important for efficacy. Discontinuation and reinitiation patterns for patients with and without type 2 diabetes (T2D) are not well-understood.

**Objective:** To describe rates and factors associated with discontinuation and reinitiation of GLP-1 RA, for patients with and without T2D.

**Design:** In this retrospective cohort study, adults with overweight or obesity initiated on GLP-1 RA between January 2018 and December 2023 were identified using electronic health record (EHR) data from a collective of 30 US healthcare systems. Patients were followed for up to 2 years to assess discontinuation and for 2 additional years to assess reinitiation.

**Setting:** Clinical and prescribing data from EHRs linked to dispensing information

**Participants:** Adults newly initiated on GLP-1 RA between 2018 and 2023, with a baseline BMI ≥27 and an available weight measurement within 60 days before initiation, and regular care in the year before initiation.

**Exposure/Covariates:** Patients were stratified by presence of T2D at baseline. Associations with socio-demographics, health factors, weight changes, and gastrointestinal (GI) adverse events (AE) were modeled.

**Main Outcomes and Measures:** Proportion of patients discontinuing and reinitiating GLP-1 RA were estimated from Kaplan-Meier models. Associations between covariates and discontinuation and reinitiation outcomes were modeled using time-varying Cox proportional hazards models.

All analyses were conducted for patients with and without T2D.

**Results:** Among 96,544 adults initiating GLP-1 RA, the mean (SD) age was 55.1 (13.3) years, 65.2% were female, 73.7% were white, and 61.3% had T2D. Individual income exceeded $50,000 for 49.7% of patients with and 57.2% of patients without T2D. One-year discontinuation was significantly higher for patients without T2D (65.1%), compared to those with T2D (45.8%). Higher weight loss, absence of GI AE, and higher income (T2D only) were significantly associated with higher discontinuation. Of 28,142 who discontinued and had a discontinuation weight available, one-year reinitiation was lower for those without T2D (34.7%), compared to those with T2D (51.0%). Weight re-gain was significantly associated with increased reinitiation.

**Conclusions and Relevance:** Most patients with overweight or obesity discontinue GLP-1 RA within 1 year, but those without T2D discontinue at higher rates and reinitiate at lower rates.

Inequities in access and adherence to effective treatments have the potential exacerbate disparities in obesity.

**Key points:** *Question:* How frequently do adults with overweight or obesity discontinue and subsequently reinitiate GLP-1 RA? What factors are associated with these outcomes?

*Findings:* In this study of 96,544 patients initiating GLP-1 RA, 46% of patients with and 65% without type 2 diabetes (T2D) discontinued within 1 year. Weight loss, income, gastrointestinal adverse events, and comorbidities were significantly associated with discontinuation. Following discontinuation, 51% of patients with and 35% without T2D reinitiated within a year. Weight re-gain since discontinuation was significantly associated with reinitiation.

*Meaning:* While most patients discontinue GLP-1 RA within a year, discontinuation is significantly higher and reinitiation is significantly lower for patients without T2D. Weight changes, tolerability, and proxies of access to care are significantly associated with sustained treatment.

## Introduction

Over 73% of US adults have obesity or overweight, ^1^ incurring approximately $173 billion in associated medical costs annually.^2^ While glucagon-like peptide-1 receptor agonists (GLP-1 RAs) substantially lower weight,^3–7^ hemoglobin A1c, and cardiovascular risk ^8^, trials suggest they must be continued for sustained effects.^9,10^

Ongoing use of GLP-1 RAs may be limited by issues related to tolerability, efficacy, access, and cost. Over 70% of patients in clinical trials reported adverse events, typically mild gastrointestinal.^3–7^ Although most patients on a GLP-1 RA achieve clinically meaningful weight loss, heterogeneity in response exists.^3–7,11^ Many patients’ weight loss also plateaus.^5,6,12^ Accordingly, a lack of desired weight loss could lead to discontinuation for some. Because obesity is more prevalent in low-income individuals, access and affordability to GLP-1 RAs may represent a major barrier to initiation and continued use.^13,14^ In addition, limited payer coverage of anti-obesity medications (AOMs) and off-label use of anti-diabetes medications (ADM), including GLP-1 RAs, still represent a major challenge for many patients.^15–17^ Finally, shortages of GLP-1 RAs remain a persistent problem.^18^

High and widely varying rates of GLP-1 RA discontinuation at 1 year, ranging from 37% to 81%, have been reported.^19–23^ Multiple factors have been found to be associated with discontinuation, including cost, insurance type, comorbidities, and absence of type 2 diabetes (T2D).^21,22^ Importantly, the magnitude of weight loss (and its effect on GLP-1 RA discontinuation) has largely been omitted from previous analyses. In addition, patterns of reinitiation following discontinuation have been poorly characterized. Because reinitiation may be influenced by the same drivers as discontinuation, further insights are needed.

This study describes rates and factors associated with GLP-1 RA discontinuation in adults with and without T2D and factors associated with reinitiation of GLP-1 RAs. The role of time-varying weight change was included in both analyses.

## Methods

### Study Design

New users of GLP-1 RAs (liraglutide, injectable semaglutide, or tirzepatide) with overweight or obesity (regardless of T2D) between 2018 and 2023 were included. New users were defined as those not previously dispensed any GLP-1 RA. Patients ≥18 years of age with a baseline body mass index (BMI) ≥ 27 kg/m^2^, baseline weight available, regular interactions with the health care system in the previous year, and a prescription for a GLP-1 RA in the preceding 2 months were included (see Study Population below). Patients were followed for up to 2 years for discontinuation. Patients who discontinued their GLP-1 RA and had a weight available within 60 days before discontinuation were followed for up to 2 additional years for reinitiation of any GLP-1 RA.

### Data Source

This study used a subset of Truveta Data. Truveta provides access to continuously updated and linked electronic health record (EHR) data from a collective of 30 US health care systems. This includes data related to demographics, encounters, diagnoses, vital signs (e.g., weight, BMI, blood pressure), medication requests (prescriptions), laboratory and diagnostic tests and results (e.g., HbA1c tests and values), and procedures. Updated EHR data are provided daily to Truveta by constituent health care systems. In addition to EHR data for care delivered within Truveta constituent health care systems, medication dispensing and social drivers of health (SDOH) were made available through linked third-party data. Medication dispensing (via e-prescribing) includes fills for prescriptions written both within and outside Truveta constituent health care systems, resulting in greater observability into patients’ medication history. Medication dispense histories are updated at the time of the encounter, and include fill dates, NDC or RxNorm codes, quantity dispensed, and days of medication supplied. SDOH data include individual-level factors, including income and education.

Data are normalized into a common data model through syntactic and semantic normalization. Truveta Data are then de-identified by expert determination under the HIPAA Privacy Rule. Once de-identified, data are available for analysis in R or Python using Truveta Studio. Data for this study were accessed on May 30, 2024.

### Study Population

We identified adults with overweight or obesity first dispensed a GLP-1 RA (liraglutide, injectable semaglutide, or tirzepatide) between January 2018 and December 2023. We required a BMI ≥ 27kg/m^2^ and a weight measured in the 60 days preceding the first dispense of GLP-1 RA. To improve outcome observability, we restricted the analysis to patients receiving usual care within a Truveta health care system in the prior year, defined as an interaction in both consecutive six-month periods in the year before GLP-1 RA initiation. Patients with a history of type 1 diabetes, gestational diabetes, or diabetic retinopathy, as well as those with missing gender or no follow-up encounters following GLP-1 RA initiation were excluded. Reinitiation analyses were further restricted to patients who discontinued GLP-1 RA within 2 years, for whom there was an encounter following GLP-1 RA discontinuation and a weight available up to 60 days before to one day after discontinuation of the GLP-1 RA.

The first GLP-1 RA dispense was considered the initiation date and the first GLP-1 RA dispensed was considered the index medication. Brand names were used to classify index medications as AOMs or ADMs. Patients were classified as having T2D if they had a diagnosis code for T2D, used insulin or DPP-4, or had an A1c > 7.5 in the 2 years before GLP-1 RA initiation.

### Outcomes

Outcomes of primary discontinuation and reinitiation were considered separately. Primary discontinuation was defined as the first date a patient was ≥60 days without any GLP-1 RA on hand, using fill dates and days of medication supplied to account for excess (carryover) medication from previous fills. Fills of all GLP-1 RAs were considered; GLP-1 RAs dispensed on subsequent fills were not required to match the index GLP-1 RA brand. Reinitiation was defined as the first fill of any GLP-1 RA following primary discontinuation.

Both discontinuation and reinitiation were treated as time-to-event outcomes to account for censoring. Patients were followed until the first outcome occurrence, the last encounter before study end (May 30, 2024), or 2 years from the index event. In discontinuation analyses, the first GLP-1 RA dispense served as the index event. In reinitiation analyses, the first discontinuation date served as the index event.

### Associations with Discontinuation

Associations between discontinuation and factors related to safety, tolerability, efficacy, health factors, socio-demographics, and access were considered. Safety and tolerability were modeled as on-treatment time-varying presence of a moderate or severe GI adverse event (bowel obstruction, cholecystitis, cholelithiasis, gastroenteritis, gastroparesis, or pancreatitis). Mild AEs, such as nausea and vomiting, were not included, given the expectation of inconsistent EHR capture. Efficacy was modeled as on-treatment time-varying change in weight relative to baseline, where baseline was the most recent weight in the 60 days before initiation. For interpretability, this was modeled per 1% weight loss. Weight change values were updated each time a new weight value was available. Weight cleaning approaches have been described elsewhere.^11^ Health factors included baseline BMI along with presence of chronic kidney disease (CKD) and/or heart failure (HF) at baseline. Separate models were used for patients with and without T2D given the expectation of different relationships between covariates and discontinuation outcomes. Sociodemographic and access factors included sex, age, race, ethnicity, age ≥65 years (a proxy for Medicare insurance), and individual income. Insurance status was not available. Fixed effects for year of initiation were included to adjust for differences in marginal discontinuation over time.

### Associations with Reinitiation

For those who discontinued their GLP-1 RA, reinitiation analyses considered the same factors as for discontinuation, but used time invariant variables for on-treatment presence of any GI AE and total on-treatment weight change (change from baseline to discontinuation). Time-varying weight change since discontinuation was also included to consider the potential impact of weight re-gain since the GLP-1 RA was stopped. For interpretability, this was modeled per 1% weight gain since discontinuation. Weight change values were updated each time a new weight value was available, with changes reported relative to the discontinuation weight.

### Statistical Analysis

The proportion of at-risk patients that discontinued and reinitiated treatment within 2 years were extracted from separate Kaplan-Meier (KM) models stratified by presence of T2D. A sensitivity analysis was performed to evaluate patients for whom 2 years of follow-up were fully available. Associations between potential drivers and outcomes of discontinuation and reinitiation for those with and without T2D were estimated using separate time-varying Cox proportional hazards models inclusive of the variables described above.

## Results

Overall, 96,544 adults met inclusion criteria, including 59,268 patients with T2D (58,551 [98.8%] using ADM and 717 [1.2%] using AOM) and 37,276 patients without T2D (24,374 [65.4%] using ADM off-label and 12,902 (34.6%) using AOM). The mean (SD) age overall was 55.1 (13.3) years, 65.2% of patients were female, and 73.7% were white (Table 1a). Individual income exceeded $50,000 for 49.7% of patients with T2D and 57.2% of patients without T2D. The median (IQR) BMI at baseline was 37.3 kg/m^2^ (33.1, 42.8) (37.2 [32.9, 42.6] for patients with T2D and 37.6 [33.4, 43.0] for those without T2D), with 37% of patients classified as having class 3 obesity (BMI ≥40) at baseline.

**Table 1:** Patient characteristics. Table 1(a) contains the characteristics of initiated patients meeting study criteria, who were included in analyses of discontinuation. Table 1(b) contains the characteristics of discontinued patients meeting additional study criteria, who were included in analyses of reinitiation.

(a) Characteristics of initiated patients (discontinuation analysis)
| Characteristic | Overall<br>N = 96,544 | T2D<br>N = 59,268 | No T2D<br>N = 37,276 |
| --- | --- | --- | --- |
| Age |  |  |  |
| 18-44 | 23,716 (25%) | 9,762 (16%) | 13,954 (37%) |
| 45-64 | 50,784 (53%) | 32,577 (55%) | 18,207 (49%) |
| 65+ | 22,044 (23%) | 16,929 (29%) | 5,115 (14%) |
| Female | 62,956 (65%) | 33,658 (57%) | 29,298 (79%) |
| Race |  |  |  |
| Asian | 2,186 (2%) | 1,495 (3%) | 691 (2%) |
| Black* | 13,099 (14%) | 8,118 (14%) | 4,981 (13%) |
| White | 71,157 (74%) | 43,111 (73%) | 28,046 (75%) |
| Other | 10,102 (10%) | 6,544 (11%) | 3,558 (10%) |
| Ethnicity |  |  |  |
| Other | 4,918 (5%) | 3,177 (5%) | 1,741 (5%) |
| Hispanic or Latino | 12,724 (13%) | 8,332 (14%) | 4,392 (12%) |
| Not Hispanic or Latino | 78,902 (82%) | 47,759 (81%) | 31,143 (84%) |
| Education: Some college on record | 47,190 (50%) | 24,225 (42%) | 22,965 (63%) |
| Estimated income |  |  |  |
| 0 - 30,000 | 6,628 (7%) | 4,140 (7%) | 2,488 (7%) |
| 30,001 - 50,000 | 36,271 (38%) | 23,709 (40%) | 12,562 (34%) |
| 50,001 - 80,000 | 36,210 (38%) | 21,681 (37%) | 14,529 (39%) |
| More than 80K | 14,554 (15%) | 7,774 (13%) | 6,780 (18%) |
| Unknown | 2,881 (3%) | 1,964 (3%) | 917 (2%) |
| GLP-1 RA initiation year |  |  |  |
| 2018 | 2,900 (3%) | 2,419 (4%) | 481 (1%) |
| 2019 | 5,542 (6%) | 4,560 (8%) | 982 (3%) |
| 2020 | 4,506 (5%) | 3,467 (6%) | 1,039 (3%) |
| 2021 | 9,945 (10%) | 6,640 (11%) | 3,305 (9%) |
| 2022 | 26,549 (27%) | 14,116 (24%) | 12,433 (33%) |
| 2023 | 47,102 (49%) | 28,066 (47%) | 19,036 (51%) |
| Label |  |  |  |
| ADM | 82,925 (86%) | 58,551 (99%) | 24,374 (65%) |
| AOM | 13,619 (14%) | 717 (1%) | 12,902 (35%) |
| Baseline BMI | 37.3 (33.1, 42.8) | 37.2 (32.9, 42.6) | 37.6 (33.4, 43.0) |
| Baseline BMI class |  |  |  |
| Overweight 27-30 | 8,736 (9%) | 5,995 (10%) | 2,741 (7%) |
| Obesity Class 1 | 26,466 (27%) | 16,365 (28%) | 10,101 (27%) |
| Obesity Class 2 | 25,978 (27%) | 15,702 (26%) | 10,276 (28%) |
| Obesity Class 3 | 35,364 (37%) | 21,206 (36%) | 14,158 (38%) |
| Weight (kg) at initiation | 106.2 (91.8, 123.8) | 107.0 (92.4, | 105.2 (90.8, 122.5) |
| Comorbidities |  |  |  |
| Atrial fibrillation | 5,766 (6%) | 4,333 (7%) | 1,433 (4%) |
| Asthma | 18,429 (19%) | 11,266 (19%) | 7,163 (19%) |
| COPD | 6,096 (6%) | 4,781 (8%) | 1,315 (4%) |
| CKD | 17,417 (18%) | 14,330 (24%) | 3,087 (8%) |
| Glaucoma | 1,817 (2%) | 1,384 (2%) | 433 (1%) |
| Heart failure | 6,675 (7%) | 5,412 (9%) | 1,263 (3%) |
| Hypertension | 66,037 (68%) | 47,467 (80%) | 18,570 (50%) |
| Hyperlipidemia | 67,366 (70%) | 48,439 (82%) | 18,927 (51%) |
| Ischemic heart disease | 6,574 (7%) | 5,386 (9%) | 1,188 (3%) |
| Major depressive disorder | 21,427 (22%) | 12,767 (22%) | 8,660 (23%) |
| Medications |  |  |  |
| Metformin | 54,669 (57%) | 46,830 (79%) | 7,839 (21%) |
| SGLT2 | 17,132 (18%) | 16,110 (27%) | 1,022 (3%) |
| Insulin | 5,469 (6%) | 5,469 (9%) | 0 (0%) |
| DPP4 | 10,602 (11%) | 10,602 (18%) | 0 (0%) |
| Sulfonylurea | 18,460 (19%) | 17,920 (30%) | 540 (1%) |
| Orlistat | 229 (0%) | 70 (0%) | 159 (0%) |
| Phentermine Topiramate | 801 (1%) | 175 (0%) | 626 %) |

**(b) Characteristics of discontinued patients (reinitiation analysis)**
| Characteristic | Overall<br>N = 28,142 | T2D<br>N = 15,162 | No T2D<br>N = 12,980 |
| --- | --- | --- | --- |
| Age |  |  |  |
| 18-44 | 7,277 (26%) | 2,347 (15%) | 4,930 (38%) |
| 45-64 | 13,517 (48%) | 7,483 (49%) | 6,034 (46%) |
| 65+ | 7,348 (26%) | 5,332 (35%) | 2,016 (16%) |
| Female | 19,400 (69%) | 8,923 (59%) | 10,477 (81%) |
| Race |  |  |  |
| Asian | 489 (2%) | 283 (2%) | 206 (2%) |
| Black* | 3,916 (14%) | 2,114 (14%) | 1,802 (14%) |
| White | 20,908 (74%) | 11,147 (74%) | 9,761 (75%) |
| Other | 2,829 (10%) | 1,618 (11%) | 1,211 (9%) |
| Ethnicity |  |  |  |
| Other | 3,721 (13%) | 2,101 (14%) | 1,620 (12%) |
| Hispanic or Latino | 23,093 (82%) | 12,272 (81%) | 10,821 (83%) |
| Not Hispanic or Latino | 1,328 (5%) | 789 (5%) | 539 (4%) |
| Education: Some college on record | 13,377 (48%) | 7,876 (61%) | 5,501 (36%) |
| Estimated income |  |  |  |
| 0 - 30,000 | 2,283 (8%) | 1,295 (9%) | 988 (8%) |
| 30,001 - 50,000 | 11,167 (40%) | 6,492 (43%) | 4,675 (36%) |
| 50,001 - 80,000 | 10,000 (36%) | 5,148 (34%) | 4,852 (37%) |
| More than 80K | 3,810 (14%) | 1,648 (11%) | 2,162 (17%) |
| Unknown | 882 (3%) | 579 (4%) | 303 (2%) |
| GLP-1 RA initiation year |  |  |  |
| 2018 | 1,144 (4%) | 944 (6%) | 200 (2%) |
| 2019 | 1,808 (6%) | 1,468 (10%) | 340 (3%) |
| 2020 | 1,402 (5%) | 1,033 (7%) | 369 (3%) |
| 2021 | 3,585 (13%) | 2,261 (15%) | 1,324 (10%) |
| 2022 | 8,807 (31%) | 4,009 (26%) | 4,798 (37%) |
| 2023 | 11,396 (40%) | 5,447 (36%) | 5,949 (46%) |
| Label |  |  |  |
| ADM | 23,403 (83%) | 14,873 (98%) | 8,530 (66%) |
| AOM | 4,739 (17%) | 289 (2%) | 4,450 (34%) |
| Baseline BMI | 37.5 (33.1, 43.1) | 37.0 (32.7, 42.7) | 37.9 (33.6, 43.4) |
| Baseline BMI class |  |  |  |
| Overweight 27-30 | 2,537 (9%) | 1,635 (11%) | 902 (7%) |
| Obesity Class 1 | 7,576 (27%) | 4,177 (28%) | 3,399 (26%) |
| Obesity Class 2 | 7,481 (27%) | 3,947 (26%) | 3,534 (27%) |
| Obesity Class 3 | 10,548 (37%) | 5,403 (36%) | 5,145 (40%) |
| Weight (kg) at initiation | 105.8 (91.2, 123.4) | 105.5 (90.7, | 106.1 (91.6, 123.7) |
| Weight (kg) at discontinuation | 101.8 (87.1, 119.3) | 100.5 (85.5, | 103.0 (88.5, 120.2) |
| Weight loss % on treatment | 2.7 (0.0, 6.6) | 2.2 (-0.2, 5.5) | 3.5 (0.3, 8.0) |
| Duration (days) on treatment | 162 (105, 277) | 162 (116, 276) | 162 (102, 277) |
| GI AE on treatment | 1,109 (4%) | 674 (4%) | 435 (3%) |
| Comorbidities |  |  |  |
| Atrial fibrillation | 1,917 (7%) | 1,356 (9%) | 561 (4%) |
| Asthma | 5,972 (21%) | 3,217 (21%) | 2,755 (21%) |
| COPD | 2,173 (8%) | 1,628 (11%) | 545 (4%) |
| CKD | 5,725 (20%) | 4,502 (30%) | 1,223 (9%) |
| Glaucoma | 580 (2%) | 422 (3%) | 158 (1%) |
| Heart failure | 2,419 (9%) | 1,892 (12%) | 527 (4%) |
| Hypertension | 19,269 (68%) | 12,482 (82%) | 6,787 (52%) |
| Hyperlipidemia | 19,258 (68%) | 12,529 (83%) | 6,729 (52%) |
| Ischemic heart disease | 2,220 (8%) | 1,755 (12%) | 465 (4%) |
| Major depressive disorder | 7,175 (25%) | 3,720 (25%) | 3,455 (27%) |
| Medications |  |  |  |
| Metformin | 14,033 (50%) | 11,589 (76%) | 2,444 (19%) |
| SGLT2 | 4,502 (16%) | 4,209 (28%) | 293 (2%) |
| Sulfonylurea | 5,155 (18%) | 5,004 (33%) | 151 (1%) |

The median (IQR) time between GLP-1 RA initiation and censoring (last encounter before May 30, 2024 or 2 years, whichever occurred first) was 484 (333, 730) days. Overall, 66,122 (68.5%) patients were observed for at least 1 year and 26,633 (27.6%) for at least 2 years following GLP-1 RA initiation.

### Discontinuation

In total, 56,705 patients discontinued their GLP-1 RA within 2 years. Accounting for censoring, 53.4% (95% CI: 53.0% - 53.7%) discontinued by 1 year and 71.8% (95% CI: 71.5% - 72.2%) discontinued by 2 years. Discontinuation rates were significantly lower for patients with T2D (45.8% [45.4% - 46.2%] by 1 year and 63.6% [63.0% - 64.1%] by 2 years) compared to patients without T2D (65.1% [64.5% - 65.6%] by 1 year and 84.9% [84.3% - 85.4%] by 2 years) (Figure 1a). Similar discontinuation rates were observed in a sensitivity analysis limited to those followed for 2 full years (n = 26,633; 54.1% [53.5% - 54.7%] discontinued by 1 year and 70.8% [70.3% - 71.4%] by 2 years).

**Figure 1:**
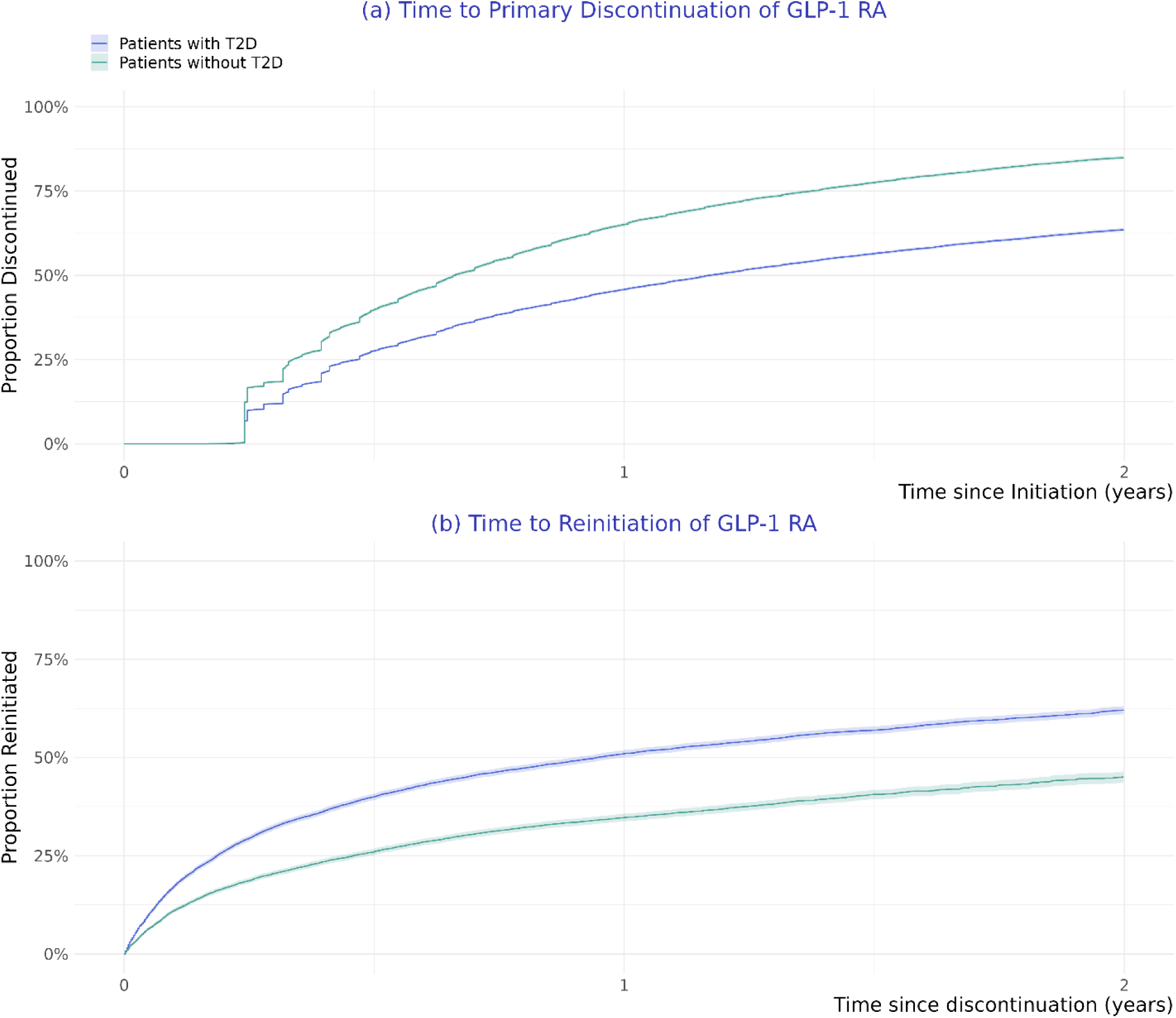
Proportions discontinuing and reinitiating GLP-1 RA. Discontinuation is the first date a patient is ≥60 days without any GLP-1 medication on hand. Reinitiation is the first fill of any GLP-1 following discontinuation. Patients were censored at their last encounter before May 30, 2023 or administrative censoring (2 years), whichever occurred first. Y-axis represents the event probability (1-survival probability).

Several factors were significantly associated with discontinuation (Figure 2). Patients ≥65 years (a proxy for Medicare eligibility) were more likely to discontinue their GLP-1 RA both with and without T2D, respectively (HR [95% CI]: 1.3 [1.3 - 1.3] and 1.1 [1.1 - 1.2]). Higher income was strongly and progressively associated with a lower rate of discontinuation in those with T2D. Compared to those with incomes under $30,000, hazards of discontinuation were 0.9 (0.8 - 0.9), 0.8 (0.8 - 0.8), and 0.7 (0.7 - 0.8) for those with incomes of $30,001 - $50,000, 50,001 - 80,000, and >$80,000, respectively. Higher on-treatment weight loss was also significantly associated with lower discontinuation. Each 1% reduction in weight from baseline was associated with a 2.9% (2.7% - 3.1%) and 3.2% (3.1% - 3.4%) lower hazard of discontinuation for patients with and without T2D, respectively. Finally, on-treatment moderate or severe incident GI AEs were associated with a higher hazard of discontinuation for patients with and without T2D, respectively (HR: 1.3 [1.2 - 1.4] and 1.2 [1.1 - 1.3]).

**Figure 2:**
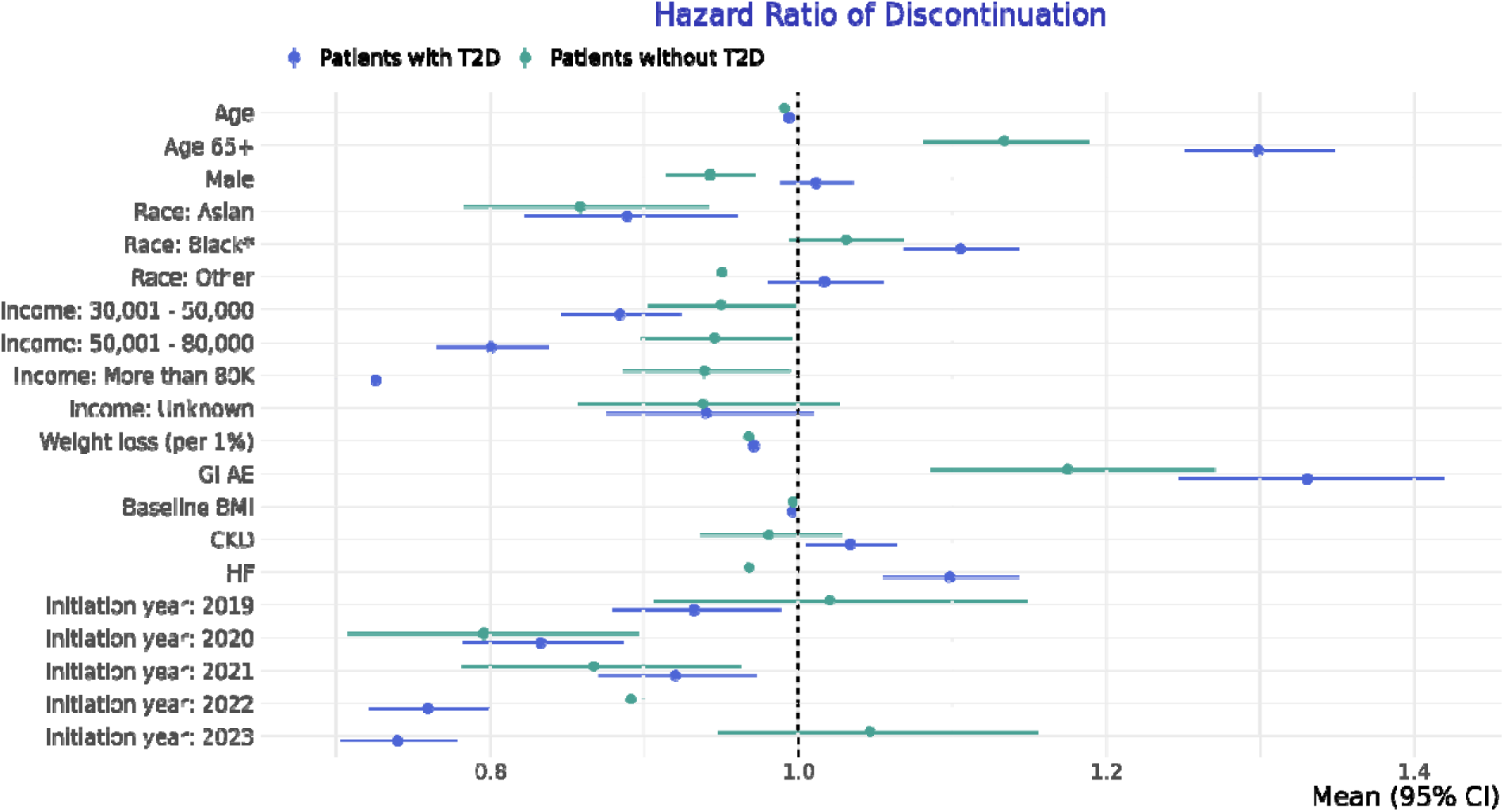
Associations (hazard ratios) between covariates and discontinuation outcomes for patients with and without T2D. Points represent hazard ratios from separate Cox proportional hazards models for patients with and without T2D. Lines represent 95% confidence intervals.

### Reinitiation

Among 56,705 patients who discontinued their GLP-1 RA, 28,142 (49.6%) were included for reinitiation analyses based on having a weight recorded at discontinuation and ≥1 encounter after discontinuation. The median (IQR) duration of primary treatment with a GLP-1 RA was 162 (105, 277) days, with a median (IQR) on-treatment weight change of -2.7% (-6.6%, 0.0%).

Similar to those who initiated a GLP-1 RA, the population considered for reinitiation was mostly female, white, and not Hispanic/Latino (Table 1b). Following primary discontinuation, the median (IQR) follow-up time was 334 (167, 667) days, with 45.0% followed at least 1 year and 22.3% followed at least 2 years.

Accounting for censoring, 51.0% (50.1% - 51.9%) of patients with T2D and 34.7% (33.8% - 35.7%) of patients without T2D reinitiated a GLP-1 RA within 1 year and 62.1% (61% - 63.1%) with T2D and 45.0% (43.6% - 46.5%) without T2D reinitiated a GLP-1 RA within 2 years of discontinuation (Figure 1b).

Several factors were significantly associated with reinitiation (Figure 3). Those ≥65 years, a proxy for Medicare eligibility, were significantly less likely to reinitiate their GLP-1 RA both with and without T2D (HR: 0.9 (0.8 - 0.9) and 0.8 [0.7 - 0.9]). While relationships between income and reinitiation were not statistically significant in either group, point estimates suggested a positive correlation between higher income and higher reinitiation. In addition, each 1% weight gain since discontinuation was significantly associated with a 2.0% (1.5% - 2.5%) and 2.2% (1.5% - 2.9%) increased hazard of reinitiation for patients with and without T2D, respectively. Finally, moderate or severe GI AEs during initial treatment were associated with a lower rate of reinitiation for patients with and without T2D, respectively (HR: 0.8 [0.7 - 0.9] and 0.7 [0.6 - 0.9]).

**Figure 3:**
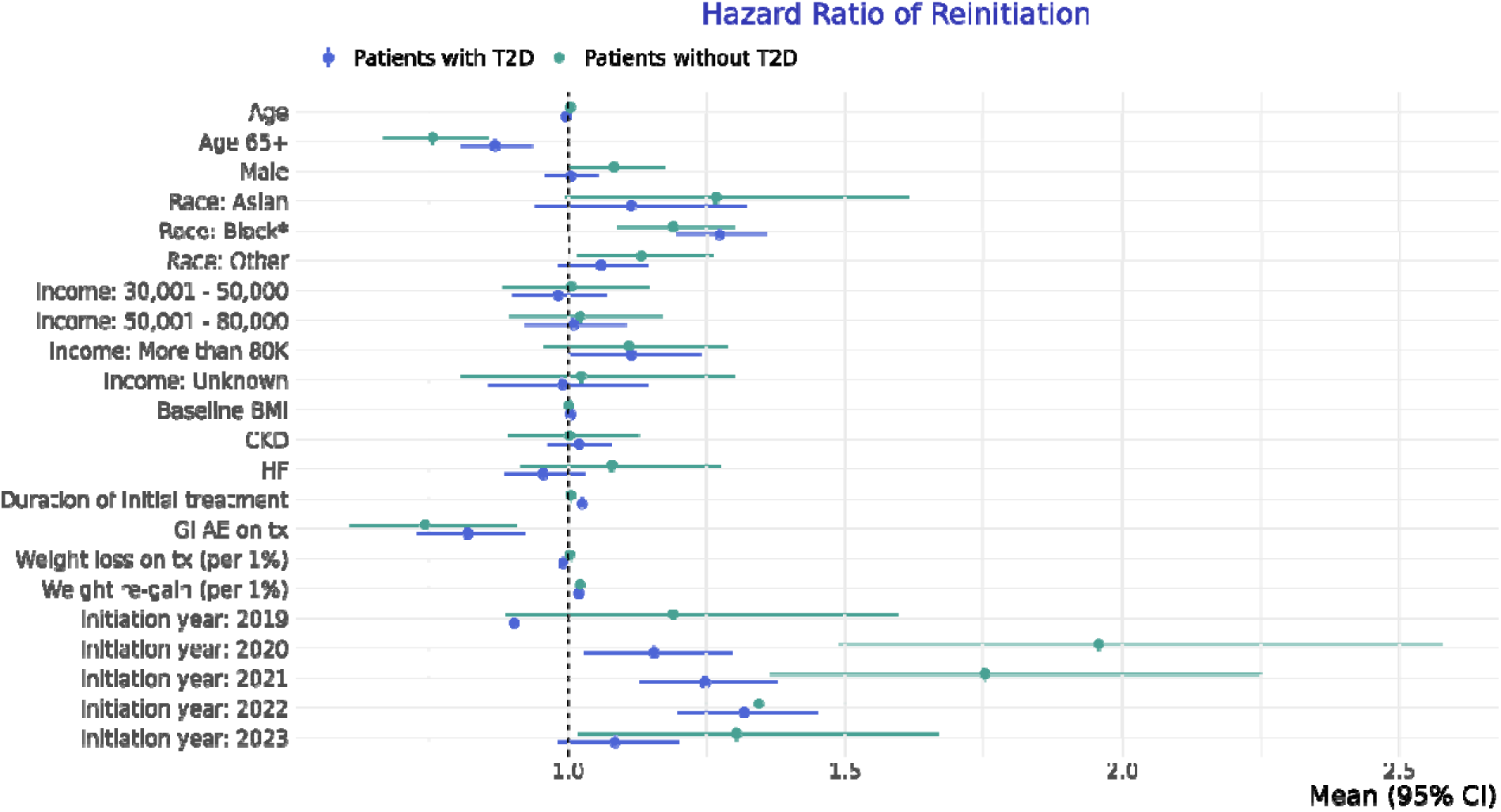
Associations (hazard ratios) between covariates and reinitiation outcomes for patients with and without T2D. Points represent hazard ratios from separate Cox proportional hazards models for patients with and without T2D. Lines represent 95% confidence intervals.

## Discussion

Most patients newly initiated on a GLP-1 RA discontinued therapy within 1 year, with a significantly higher discontinuation rate among patients without T2D. These same non-T2D patients also had a significantly lower rate of GLP-1 RA reinitiation. Greater insurance coverage of GLP-1 RA for patients with T2D may explain these differences in discontinuation and reinitiation. Greater on-treatment weight loss was associated with a lower likelihood of GLP-1 RA discontinuation; similarly, weight gain since discontinuation was associated with an increased likelihood of GLP-1 RA reinitiation. Three points need emphasis.

First, the high rate of GLP-1 RA discontinuation observed in this study (53.4% by one year) is consistent with that found in other recent analyses. Recent studies have found discontinuation rates ranging from 37-81%, depending on the population, data source, and time period considered.^19–24^ Importantly, the finding of an increased discontinuation rate for GLP-1 RAs among patients without T2D and those with heart failure and adverse GI AEs is similar to that noted in previous studies.^22^

Second, the influence of weight loss on discontinuation and weight re-gain on reinitiation suggests that weight management is an important factor regardless of T2D status.

Finally, lower income was strongly associated with higher discontinuation in patients with T2D, but not those without T2D. Patients without T2D had higher incomes so the cost of GLP-1 RAs may have been less important in their decisions. Furthermore, patients highly sensitive to affordability may have declined to initiate GLP-1 in the first place, given limited insurance coverage for patients without T2D. Additionally, lower-cost treatment alternatives, like metformin or DPP-4, are available for patients with T2D.^25^ In contrast, the lack of effective alternative treatments for patients without T2D may necessitate GLP-1 RA treatment regardless of cost. Further studies should investigate the contribution of insurance coverage and out-of-pocket costs on GLP-1 RA adherence.

Our findings have important policy implications related to equitable access and outcomes for patients with and without T2D. Lack of drug adherence may limit the long-term health benefits of GLP-1 RA, such as cardiovascular risk reduction. Our analysis suggests greater efforts are needed to increase access and adherence for patients without T2D and those with lower incomes.

This study has several strengths. To our best knowledge, this is the first study to characterize patterns of reinitiation following GLP-1 RA discontinuation. Use of EHR data also allowed for inclusion of weight changes during treatment and after discontinuation. Given limited insurance coverage of AOMs, use of linked EHR and dispensing information allowed inclusion of patients paying for GLP-1 RAs out-of-pocket. Lastly, use of survival models, instead of binary outcomes models, allowed for use of very recent data that was subject to high censoring.

Importantly, this study also has several limitations. First, GLP-1 RA shortages were not modeled explicitly in this analysis given insufficient data. While inclusion of fixed effects for the year of initiation may account for some differences in medication availability over time, it is unlikely to fully characterize shortage-related effects. Second, discontinuation definitions vary in the literature and are likely subject to sensitivity vs. specificity tradeoffs. While we defined discontinuation as 60 days without any medication on hand based on both clinical and data considerations, some patients may have been misclassified. Furthermore, medication rationing (skipping doses) was not considered in this analysis. Finally, mild GI AEs (e.g., nausea, vomiting) were not included in the analysis, given an expectation of inconsistent capture in the EHR.

## Conclusions

Most adults with overweight or obesity initiated on a GLP-1 RA discontinued therapy within 1 year. Those without T2D discontinued their GLP-1 RA at a significantly higher rate and reinitiated a GLP-1 RA at a significantly lower rate, compared to those with T2D. Greater weight loss and higher income were related to lower discontinuation, while weight re-gain since discontinuation was associated with higher reinitiation. Access to and insurance coverage of GLP-1 RAs for patients without T2D may contribute to these differences.

## Data Availability

The data used in this study are available to all Truveta subscribers and may be accessed at studio.truveta.com.

## Notes

### Competing Interest Statement

All authors except EJE, VZ, and TJG are employees of Truveta, Inc.

### Funding Statement

This study did not receive any funding.

### Author Declarations

Normalized electronic health record data are de-identified by expert determination under the HIPAA Privacy Rule before being made available to researchers. In accordance with 45 C.F.R. Para. 46.101 Protection of Human Subjects, our study did not require Institutional Review Board approval because it used only deidentified medical records. All data used in this study are publicly available to Truveta subscribers and may be accessed at studio.truveta.com.

## References

1. Fryar CD, Carroll MD, Afful J. Prevalence of overweight, obesity, and severe obesity among adults aged 20 and over: United States, 1960–1962 through 2017–2018. NCHS Health E-Stats. 2020. Internet: https://www.cdc.gov/nchs/data/hestat/obesity-adult-17-18/obesity-adult.htm (accessed 24 August 2021). Published online 2022.

2. Ward ZJ, Bleich SN, Long MW, Gortmaker SL. Association of body mass index with health care expenditures in the United States by age and sex. PLOS ONE. 2021;16(3):e0247307. doi:10.1371/journal.pone.0247307

3. Davies M, Færch L, Jeppesen OK, et al. Semaglutide 2· 4 mg once a week in adults with overweight or obesity, and type 2 diabetes (STEP 2): a randomised, double-blind, double-dummy, placebo-controlled, phase 3 trial. The Lancet. 2021;397(10278):971–984.

4. Garvey WT, Frias JP, Jastreboff AM, et al. Tirzepatide once weekly for the treatment of obesity in people with type 2 diabetes (SURMOUNT-2): a double-blind, randomised, multicentre, placebo-controlled, phase 3 trial. The Lancet. Published online 2023.

5. Jastreboff AM, Aronne LJ, Ahmad NN, et al. Tirzepatide Once Weekly for the Treatment of Obesity. New England Journal of Medicine. 2022;387(3):205–216. doi:10.1056/NEJMoa2206038

6. Wilding JPH, Batterham RL, Calanna S, et al. Once-Weekly Semaglutide in Adults with Overweight or Obesity. New England Journal of Medicine. 2021;384(11):989–1002. doi:10.1056/NEJMoa2032183

7. Davies MJ, Bergenstal R, Bode B, et al. Efficacy of Liraglutide for Weight Loss Among Patients With Type 2 Diabetes: The SCALE Diabetes Randomized Clinical Trial. JAMA. 2015;314(7):687–699. doi:10.1001/jama.2015.9676

8. Lincoff AM, Brown-Frandsen K, Colhoun HM, et al. Semaglutide and Cardiovascular Outcomes in Obesity without Diabetes. New England Journal of Medicine. 2023;389(24):2221–2232. doi:10.1056/NEJMoa2307563

9. Aronne LJ, Sattar N, Horn DB, et al. Continued Treatment With Tirzepatide for Maintenance of Weight Reduction in Adults With Obesity: The SURMOUNT-4 Randomized Clinical Trial. JAMA. 2024;331(1):38–48. doi:10.1001/jama.2023.24945

10. Wilding JPH, Batterham RL, Davies M, et al. Weight regain and cardiometabolic effects after withdrawal of semaglutide: The STEP 1 trial extension. doi:10.1111/dom.14725

11. Rodriguez PJ, Goodwin Cartwright BM, Gratzl S, et al. Semaglutide vs Tirzepatide for Weight Loss in Adults With Overweight or Obesity. JAMA Internal Medicine. Published online July 8, 2024. doi:10.1001/jamainternmed.2024.2525

12. Hall KD. Physiology of the weight-loss plateau in response to diet restriction, GLP-1 receptor agonism, and bariatric surgery. Obesity. 2024;32(6):1163–1168. doi:10.1002/oby.24027

13. Ogden CL, Fakhouri TH, Carroll MD, et al. Prevalence of Obesity Among Adults, by Household Income and Education — United States, 2011–2014. MMWR Morb Mortal Wkly Rep. 2017;66(50):1369–1373. doi:10.15585/mmwr.mm6650a1

14. Wright DR, Guo J, Hernandez I. A Prescription for Achieving Equitable Access to Antiobesity Medications. JAMA Health Forum. 2023;4(4):e230493. doi:10.1001/jamahealthforum.2023.0493

15. Baig K, Dusetzina SB, Kim DD, Leech AA. Medicare Part D Coverage of Antiobesity Medications — Challenges and Uncertainty Ahead. N Engl J Med. 2023;388(11):961–963. doi:10.1056/NEJMp2300516

16. Vokinger KN, Nussli E, Dusetzina SB. Access to GLP-1 Weight Loss Drugs in the US, Canada, Switzerland, and Germany. JAMA Internal Medicine. Published online July 15, 2024. doi:10.1001/jamainternmed.2024.2559

17. Liu BY, Rome BN. State Coverage and Reimbursement of Antiobesity Medications in Medicaid. JAMA. 2024;331(14):1230–1232. doi:10.1001/jama.2024.3073

18. Whitley HP, Trujillo JM, Neumiller JJ. Potential Strategies for Addressing GLP-1 and Dual GLP-1/GIP Receptor Agonist Shortages. Clinical Diabetes. Published online 2023:cd230023.

19. Durden E, Liang M, Fowler R, Panton UH, Mocevic E. The Effect of Early Response to GLP-1 RA Therapy on Long-Term Adherence and Persistence Among Type 2 Diabetes Patients in the United States. Journal of Managed Care &amp\mathsemicolon Specialty Pharmacy. 2019;25(6):669–680. doi:10.18553/jmcp.2019.18429

20. Weiss T, Carr RD, Pal S, et al. Real-world adherence and discontinuation of glucagon-like peptide-1 receptor agonists therapy in type 2 diabetes mellitus patients in the United States. Patient preference and adherence. Published online 2020:2337–2345.

21. Gasoyan H, Pfoh ER, Schulte R, Le P, Rothberg MB. Early and LATER STAGE persistence with ANTIOBESITY medications: A retrospective cohort study. Obesity. 2024;32(3):486–493. doi:10.1002/oby.23952

22. Do D, Lee T, Peasah SK, Good CB, Inneh A, Patel U. GLP-1 Receptor Agonist Discontinuation Among Patients With Obesity and/or Type 2 Diabetes. JAMA Network Open. 2024;7(5):e2413172–e2413172.

23. Gleason PP, Urick BY, Marshall LZ, Friedlander N, Qiu Y, Leslie RS. Real-world persistence and adherence to glucagon-like peptide-1 receptor agonists among obese commercially insured adults without diabetes. JMCP. Published online May 8, 2024:1–8. doi:10.18553/jmcp.2024.23332

24. Real-World Trends in GLP-1 Treatment Persistence and Prescribing for Weight Management. Blue Health Intelligence (BHI) for Blue Cross Blue Shield Association

25. Quach J, Midlam C, Sell C, Levin-Scherz J. Out-of-Pocket Costs for Diabetes Medications in Employer-Sponsored Health Insurance Plans. 2024;30:107–108.

